# Effectiveness of ReCOVery APP to improve the quality of life of Long COVID patients: a 6-month follow-up randomized clinical trial

**DOI:** 10.1101/2023.08.30.23294831

**Authors:** Mario Samper-Pardo, Bárbara Oliván-Blázquez, Sandra León-Herrera, Rafael Sánchez-Arizcuren, Verónica Casado-Vicente, Raquel Sánchez-Recio

## Abstract

The main objective of this study is to analyse the clinical efficacy of medium-term telerehabilitation in the recovery of patients with Long COVID using ReCOVery APP, administered in the Primary Health Care (PHC) setting. The second objective is to identify significant patterns associated with an improvement in their quality of life predicted by other study variables. To this end, a randomised clinical trial was conducted with two parallel groups of a total of 100 patients with Long COVID. The control group continued with their usual treatment (TAU), established by their primary care physician. The intervention group, in addition to continuing with their TAU, attended three sessions based on motivational methodology and used ReCOVery APP for six months. The main variable was quality of life. The results of this study concluded that ReCOVery APP was not significantly more effective in improving the quality of life of patients with Long COVID. There was low adherence of participants. However, linear regression analyses revealed significant patterns of improvement in overall quality of life and mental health predicted by time of use of the APP and the personal construct of self-efficacy. In addition, all participants significantly improved their physical and mental health over the duration of the intervention. In conclusion, meaningful use of the ReCOVery APP may contribute to improving the quality of life of patients with Long COVID, but strategies to improve adherence need to be encouraged.

**Trial Registration No:** ISRCTN91104012.

## INTRODUCTION

The severe and global impact of COVID-19 has generated a pandemic of consequences. Among them, since 2020 many infected people continue to have persistent and debilitating symptoms for months after infection (1). It was in October 2021 when the World Health Organization (WHO) defined these persistent symptoms as a new pathology, calling it as Post COVID-19 Condition (2). It consists of symptoms typical of a probable or confirmed SARS-CoV-2 infection that continue or develop three months after the infection, and that do not correspond to an alternative diagnosis. The scientific society refers to this pathology as "Long COVID" or "persistent COVID", among others.

According to the WHO (3), approximately 145 million people have been affected by Long COVID during the first two years of the pandemic worldwide. The etiology of Long COVID is still a great unknown, including its origin, duration, or possible treatments. This fact represents a gap for the society of the moment, especially for the scientific and health community. Emerging evidence seems to indicate a high prevalence of persistent COVID among people hospitalized for COVID-19 (4). In fact, Ceban et al., (2022) estimated this prevalence of 10-30% of non-hospitalized COVID-19 patients, compared to 50-70% in hospitalized COVID-19 patients (5). Also, this high prevalence has been reflected in the female sex and in ages around 50 years, although often based on cross-sectional studies of small samples (6). More recently, several hypotheses state that comorbidities (7) and the immune response (8) could be behind the development of Long COVID illnesses.

The symptoms of persistent COVID can affect multiple organ systems (9), mainly: respiratory, cardiovascular, gastrointestinal, neurological, musculoskeletal, dermatological, visual, and olfactory, in addition to chronic fatigue and a negative impact on their emotional well-being (10). Davis et al. (2021) has come to count 203 symptoms of this disease (11). The development and evolution of these persistent symptoms supposes a total alteration of their organism (11). They are disabling, altering their personal, family, social and work environment and, consequently, their quality of life (12).

These sequelae require multidisciplinary rehabilitation, which makes it possible to address the multisystem symptoms that these patients present (13). Along these lines, given the need to continue offering general rehabilitation during the pandemic, telerehabilitation underwent an unprecedented boost that persists to this day (14,15). Even so, prior to the COVID-19 pandemic, the benefits of telerehabilitation compared to traditional rehabilitation had already been demonstrated, not only for therapeutic benefits but also in terms of cost-effectiveness (16). In this way, telerehabilitation is positioned as a real and possible alternative to help patients with long-term or chronic diseases, such as Long COVID patients. In fact, the qualitative study by Reis et al., (2022), based on the perception of nurses, considers telerehabilitation programs a fundamental strategy for recovery programs aimed at Long COVID patients (17). A meta-analysis of randomized controlled trials conducted by Hung et al., (2022), considers telerehabilitation as an effective option to improve dyspnea or muscle strength, among others, in Long COVID patients (18). Thus, a quasi-experimental pre-post study evaluated the functional capacity of Long COVID patients after four weeks of digital physiotherapy, obtaining significant improvements (19). An online seven-weeks course aimed at "Recovery from COVID" obtained significant improvements in the quality of life in the seventy-six participants who completed it (20). Also, a systematic review of exercise interventions through telerehabilitation in patients with Post COVID-19 symptoms confirms their efficacy in the short and long term (21). However, all the studies mentioned above suggest the need for large-scale randomized clinical trials to be able to extrapolate their results. Therefore, it seems that the effectiveness of telerehabilitation in this group of patients still needs to be investigated.

Given the need for multidisciplinary management of patients with persistent COVID and the growing momentum for telerehabilitation, this study developed an APP called ReCOVery. It is a digital tool with rehabilitation content based on clinical guidelines and evidence (11,22–25). An RCT with two parallel groups was conducted to assess its short-term clinical effectiveness (26). However, it is necessary to know the effectiveness of this intervention in the medium term.

Therefore, the main objective is to deepen the clinical efficacy of rehabilitation (ReCOVery APP) for people with a diagnosis of Long COVID for six months, compared to the treatment as usual (TAU) administered in the context of Primary Health Care (PHC). The second objective is to identify significant models associated with an improvement in their quality of life and other study variables.

## METHODOLOGY

### Study design

This research study is an open-label randomized clinical trial (RCT) through two parallel groups of Post COVID-19 patients infected twelve weeks or more ago and with persistent symptoms. First group participants have continued to receive their TAU established by their GP of PHC (control group). Second group participants have also continued with their TAU and have also had access to ReCOVery APP, as an adjuvant treatment in the form of telerehabilitation, and to three motivational sessions to increase adherence to the APP (intervention group).

This research was carried out in the territory of Aragon, located in the northeast of Spain. This RCT was registered in February of 2022 (ISRCTN91104012). The recruitment of patients was carried out from January 2022 to March 2022. The baseline evaluation and start of the intervention were carried out in March-April 2022, the follow-up evaluation in June-July 2022 (26), and the final evaluation was carried out in October 2022. In addition, the original protocol article of this investigation was published (27).

### Sample size

The sample size was established and detailed in the protocol article of this RCT study (27). The pre-post score difference of the Short Form-36 Health Survey Questionnaire (SF-36) instrument was used, considering the value of the highest possible standard deviation (SD) and a minimum expected difference of 19.3 points for the pre-post rating. A risk of 0.05 was accepted as well as a power of 95% in a two-sided contrast, and a maximum dropout rate of 10%. The minimum required sample size was 78 subjects.

Given the demand of the potentially interested participants, the final sample size included 100 participants, 22 more than the required sample size.

### Recruitment of participants

The study population were patients with Post COVID-19 condition, over eighteen years of age, infected for twelve weeks or more and with a positive diagnostic test for COVID-19. The exclusion criteria were: not having a positive COVID-19 diagnostic test for more than the previous twelve weeks; participation in a clinical trial in the last six months; pregnancy and lactation; have a diagnosis of severe uncontrolled illness; significant risk of suicide; existing structured psychotherapeutic or rehabilitative treatment by health professionals and the presence of any medical, psychological or social problems that may significantly interfere with the study.

Patients originating from the consultation of GPs of PHC and who met the inclusion criteria were included in the study. Also, some interested patients from the association for those affected "Long COVID Aragón" were redirected to their GP to be eligible to participate if they met the established criteria.

An informative document with the study criteria was provided to the GPs. In this way, when they identified potential patients, and after obtaining their informed consent, they contacted one of the researchers to facilitate contact with the potential patient. Subsequently, the researcher contacted the probable participant again to reconfirm if they met the criteria, their interest in the study, and to resolve any possible doubts. Recruitment was carried out consecutively until reaching the estimated sample size, from January to March 2022. Prior to carrying out the baseline evaluation of the participants, it was necessary for them to offer their written consent to participate.

### Randomization and assignment and blinding of study groups

The individual randomization process was performed using an alphabetical list of participants and computer-generated blind sequence. An independent investigator, and therefore blinded, was commissioned to carry out this task. The assignment to both groups was not blind, due to the very nature of the intervention. The same independent investigator made a phone call to each participant to confirm the assigned group and to ask her not to inform third parties about her assignment. On the one hand, the participants assigned to the control group were required to continue with their TAU and not start rehabilitation-type activities that could affect the intervention. On the other hand, the participants of the intervention group were summoned in person and individually at a nearby health center. The latter were asked to bring their personal mobile device with a battery, to install the APP.

### Development and evaluation the APP and interventions

Prior to the design of the APP, a qualitative study was carried out through individual interviews and focus groups with Long COVID patients (28). Some of the main themes were persistent symptoms, identified needs or possible treatments followed. Subsequently, the available scientific evidence on health recommendations and rehabilitation for Long COVID patients was also collected (11,22–25). In this way, a human-centered design was chosen (29), a technique that seeks to solve problems and needs by understanding the users themselves, in this case Long COVID patients. A native APP with Java language was created using the Android Studio platform. The design of a native APP was chosen, instead of a hybrid one, to allow the use of the device’s own tools, such as notifications. All its contents must be graduated and personalized according to the needs and characteristics of each patient, as indicated in the available instructions. All the details about the contents and bibliographic references of ReCOVery APP were collected in the protocol article of this RCT (26,27). ReCOVery APP consists of six main modules:

- 1) Food. Recommendations based on adherence to the Mediterranean diet are provided, with the aim of supplying possible nutritional deficiencies of vitamin D, vitamin B12, B complex, folic acid and omega-3 fatty acids.
- 2) Rest and sleep. Recommendations are provided as tips that can be carried out daily, to improve the quality of sleep and rest. The need to achieve an average of between 7 and 8 hours of sleep each night is promoted, to achieve a restful and satisfactory rest.
- 3) Physical exercises. In this section, physical rehabilitation exercises are provided through graphic representations. All the contents and indications in this section were based on guidelines on the management of this disease or other pathologies with similar symptoms.
- 4) Breathing exercises. Various video tutorials on respiratory physiotherapy are provided following the indications and symptoms of these patients.
- 5) Cognitive exercises. Three levels of cognitive stimulation exercises are provided, aimed at working on cognitive skills focused on executive function, difficulty maintaining attention, decreased processing speed, verbal fluency, and short-term memory deficits.
- 6) Participation in the community. The objective is to promote participation in the local development process through different services, associations, or cultural activities, as well as groups affected by the same pathology, seeking to improve their emotional well-being.

All the participants continued with their TAU, being cared for by their GP and other PHC professionals. However, participants assigned to the intervention group also had access to ReCOVery APP for twenty-four weeks. In addition, at the beginning of the intervention, this group of patients attended three sessions based on motivational methodology, APP management, and strengthening of their personal constructs (health literacy, self-efficacy, and personal activation) in relation to their disease. All the sessions were carried out in person, based on the guidelines of Miller and Rollnick (30). The first two sessions were individual and the third group session (in groups of 8-10 participants). The individual sessions were guided by a clinical psychologist, lasting 20-30 minutes, in which the APP was installed and doubts about its use and management were resolved. The group sessions were guided by two clinical psychologists, lasting 50-60 minutes. The sessions were carried out during three consecutive weeks, so all the participants completed the sessions in the same weeks.

### Follow-up of the intervention and adverse events

Prior to the start of the intervention, the group of researchers established as possible adverse events: reinfection by COVID-19, use of emergency medical services, hospitalization, or surgical interventions, as well as any other circumstance that could affect the development of the intervention. In addition, all participants were provided with a telephone number, where they could report adverse events at any time during the study. All reported adverse events were assessed by two independent investigators, blinded to group assignment. In case of discrepancies, a third investigator would evaluate the situation. There were no adverse events other than those mentioned above or discrepancies during the intervention.

Despite being a remote and uncontrolled intervention, two follow-up calls were made, at six weeks and eighteen weeks from the start. Information on possible adverse events was also requested on these calls.

### Measures and variables

A total of three measurements were made: baseline evaluation (T0), follow-up evaluation carried out twelve weeks from the beginning (T1) and final evaluation carried out forty-two weeks from the beginning of the intervention (T2).

All the evaluations were carried out face-to-face and individually in a health center in their city for two consecutive weeks. They were asked to come with sports or comfortable clothes. In addition, it was necessary to go with prescription glasses, hearing aids or any functional element in case of need. The evaluations were carried out by two blinded researchers with experience in similar research projects. However, both were instructed to do so through theoretical and practical sessions, avoiding biases in said process.

The main study variable is quality of life. The SF-36 (31) was selected to be evaluated. This instrument measures eight dimensions of health (physical functioning, bodily pain, general health perceptions, physical role functioning, emotional role functioning, social role functioning, vitality, and emotional well-being) that are grouped into two main components: physical health and mental health, with whom we will work in the statistical analysis. The eight scales and the two components are scored from zero to one hundred, with scores above or below fifty indicating better or worse health, respectively. Cronbach’s alpha obtained in this study was 0.89.

Regarding the secondary variables, a total of eight validated scales have been selected. The official versions in Spanish of all scales were used. Moreover, an ad hoc questionnaire was designed for variables: sociodemographic, clinical and use of ReCOVery APP.

- The sociodemographic variables have been gender (man/woman/other), age, civil status (married or in couple/single, separated, divorced or widowed), education (no studies or primary studies/secondary or university studies) and occupation (employee/ unemployed/employee with temporary work disability (TWD)/retired/others).
- The clinical variables studied have been time since infection (months) and number of self-reported persistent symptoms at the time of each evaluation, using a list of thirty persistent symptoms typical of Long COVID patients according to previous literature (7,25,32,33).
- The use variable of ReCOVery APP has been the time of use of the APP, expressed in minutes. Regarding adherence to the APP, significant use was estimated from fifteen minutes a day, five days a week, for twenty-four weeks (1,800 minutes or more).

Emotional well-being, in relation to depression and anxiety, was measured using the Hospital Anxiety and Depression Scale (HADS) questionnaire (34). It includes fourteen items, each item corresponding to a four-point Likert scale, with scores ranging from zero to forty-one for its total score. Higher scores indicate more severe symptoms. Cronbach’s alpha obtained in this study was 0.93.

- Cognitive status was assessed using the Montreal Cognitive Assessment (MoCA) questionnaire (35). It assesses cognitive domains such as memory, attention, language or working memory. The maximum score obtained is 30 points, indicating mild cognitive impairment for scores below twenty-six points, the deterioration being progressive the lower the score. Cronbach’s alpha obtained in this study was 0.49.
- Physical functioning was assessed using the thirty-second Sit to Stand Test (36). The test assesses endurance at high power, speed in terms of strength, or muscular strength, recording the number of times a person can fully stand up and sit down. It has good test-retest reliability (0.84 <R< 0.92).
- Habitual physical activity was measured using the International Physical Activity Questionnaire-Short Form (IPAQ-SF) (37). The minutes’ walked score was used in the analysis of this study. It consists of seven items and records activity at four levels of intensity. Cronbach’s alpha obtained in this study was 0.71.
- The quality of sleep and rest was measured using the Insomnia Severity Index (ISI) questionnaire (38). This scale has seven items, with a Likert Scale from zero to four, and an overall score ranging from zero to twenty-eight. A higher score indicates a greater severity of insomnia. Cronbach’s alpha obtained in this study was 0.89.
- Personal constructions. Three validated questionnaires were selected to know the personal factors related to their behaviour:

a. Self-efficacy was assessed using the Self-efficacy Scale-12 (GSES-12) (39). It is a questionnaire with twelve items, with a Likert Scale from one to five. The resulting scores range from twelve to sixty, with a higher score indicating better self-efficacy. Cronbach’s alpha obtained in this study was 0.81.
b. The activation of the patient in his own health was measured using the Patient Activation Measure questionnaire (PAM) (40). This questionnaire on managing your own health contains thirteen items with a Likert Scale from one (strongly disagree) to four (strongly agree). The scores obtained range from thirteen to fifty-two, with the highest indicating better activation. Cronbach’s alpha obtained in this study was 0.78.
c. Health literacy will be measured using the Health Literacy Europe Questionnaire (HLS- EUQ16) (41). It contains sixteen elements, ranging from one to four. The resulting score ranges from sixteen to sixty-four. A higher score indicates worse health literacy. Cronbach’s alpha obtained in this study was 0.87.

### Statistical analysis

Statistical analyzes were performed using IBM SPSS Statistics version 22.0.0.0 and Microsoft Excel software. Normality of the sample distribution was assumed when there are more than 50 participants (42).

First, a descriptive analysis of the sample was made, calculating frequencies and % for the qualitative variables and mean and standard deviation for the quantitative variables. Secondly, a univariate and bivariate analysis was performed (comparison at baseline and differences in measurements at three months and six months) calculating Chi-Square for qualitative variables and T-Student for quantitative ones.

Second, to analyze the effectiveness of ReCOVery APP, a per-protocol analysis was performed, comparing the differences at baseline (T0), at three months (T1), at six months (T2) and at six months (T2-T0) between both groups using T-Student.

Third, it was examined whether there had been a significant improvement in all subjects over the six months, regardless of their allocation group. For this, baseline (T0) and post-intervention (T2) comparisons were made, using T-Student for the related samples for the ten selected scales and the number of symptoms.

Finally, to analyze the variables associated with the effectiveness of ReCOVery APP, a linear regression model was developed for SF-36 quality of life, SF-36 physical health, and SF-36 mental health as dependent variables. The time of use from the beginning to the end of the intervention and the three personal constructs (GSES-12, PAM and HLS-EUQ16) of all the participants in the control group and intervention group who completed the study by performing the tests were included as independent variables. three measurements, and a final model was obtained. In addition, a multicollinearity test was performed. Linear regression was used since the model residuals had a finite mean, constant variance, and normal distribution. However, a bootstrapping analysis was also performed with 2000 samples.

### Ethical approval

Ethical approval was granted by the Aragon Clinical Research Ethics Committee (PI21/454), complying with all the ethical standards of the recently mentioned committee. In addition, the procedures carried out for the creation of this work also complied with the Declaration of Helsinki of 1975. All the participants signed an informed consent prior to the start of the intervention. All the data obtained in this research were anonymized and will only be used for the purposes of the study. Any change or modification relevant to the research will be notified to the corresponding ethics committee.

## RESULTS

Initially, a total of 182 people were included in the study (Figure 1). 72 (39.56%) were discarded for not meeting the inclusion criteria and 10 (5.49%) refused to participate. 100 people were randomly assigned to the study, 52 participated in the intervention group and 48 in the control group. Subsequently, in the evaluation carried out at 6 months, 20 people were discarded (8 were reinfected by COVID19 and 12 refused to continue in the study). Finally participating in the evaluation of the 6 months a total of 80 people.

**Figure 1.**
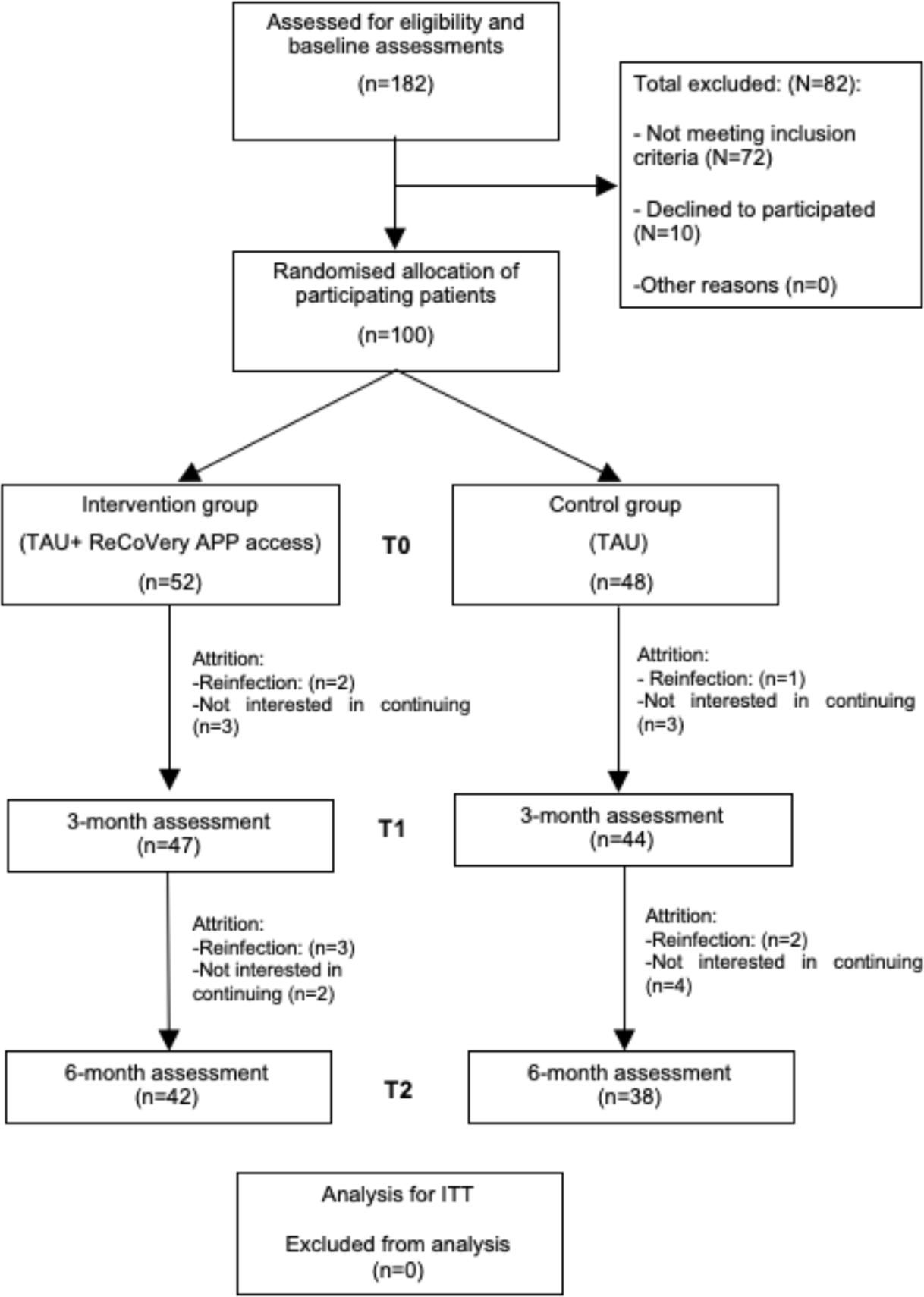
Flowchart of the study. randomization, sampling and monitoring of patients. TAU, Treatment as Usual; ITT, Intention-to-treatment

The general characteristics of the patients recruited for this study are presented in Table 1, as well as their comparison by assigned group on the variables collected at baseline. The descriptive analysis at baseline yielded a total of 100 participants (80 women and 20 men). The general profile corresponds to a woman with a mean age of 48.28 years (SD: 9.26), with secondary or university education and with active employment or TWD. The participants have a low quality of life (both physical and mental), according to the results of the SF-36. In addition, they have a mean of 16.47 (SD: 5.99) persistent symptoms. This is consistent with their limited physical functioning (reflected in the Sit to Stand Test and IPAQ-SF score), cognitive impairment (MoCA), high levels of anxious-depressive symptoms (HADS) and insomnia (ISI). In terms of personal constructs, these participants are characterised by acceptable levels of self-efficacy (GSES-12), high activation and self-management (PAM), but low levels of health literacy (HLS-EUQ16). As for the comparison by assigned group, this analysis subsequently revealed no significant differences between the groups.

**Table 1.** Description of sociodemographic and clinical variables of the total sample and comparing by assigned group.

| Variables | Total sample<br>N(%) / Median (IQR)<br>N=100 | Intervention gr.<br>N(%) / Median (IQR)<br>N=52 | Control gr.<br>N(%) / Median (IQR)<br>N=48 | P-value |
| --- | --- | --- | --- | --- |
| Gender (%) |  |  |  |  |
| Men | 20 (20%) | 8 (15,4%) | 12 | 0,230 |
| Women | 80 (80%) | 44 (84,5%) | 33 |  |
| Age | 48,28 (9,26) | 48,25 (10,36) | 48,31 (8,01) | 0,963 |
| Perceived age | 58,10 (14,69) | 59,69 (15,02) | 56,38 (14,27) | 0,260 |
| Marital status (%) |  |  |  |  |
| Married or in couple | 70 (70%) | 35 (67,3%) | 35 (72,9%) | 0,541 |
| Single, separated, widowed | 30 (30%) | 17 (32,7%) | 13 (27,1%) |  |
| Educational level (%) |  |  |  |  |
| Without studies or primary studies | 9 (9%) | 5 (9,6%) | 4 (8,3%) | 0,823 |
| Secondary or university studies | 91 (91%) | 47 (94,4%) | 44 (91,7%) |  |
| Ocupación (%) |  |  |  |  |
| Employee | 46 (46%) | 20 (38,5%) | 26 (54,2%) | 0,350 |
| Unemployed | 5 (5%) | 4 (7,7%) | 1 (2,1%) |  |
| TWD | 37 (37%) | 21 (40,4%) | 16 (33,3%) |  |
| Retired | 9 (9%) | 6 (11,5%) | 3 (6,3%) |  |
| Others | 3 (3%) | 1 (1,9%) | 2 (4,2%) |  |
| Time since the contagious | 16,12 (6,34) | 15,75 (6,56) | 16,52 (6,14) | 0,545 |
| Number of persistent symptoms | 16,47 (5,99) | 17,50 (5,29) | 15,35 (6,55) | 0,074 |
| SF-36 |  |  |  |  |
| SF-36 Physical Health | 32,19 (16,61) | 29,06 (13,67) | 35,58 (18,86) | 0,053 |
| SF-36 Mental health | 34,77(19,31) | 32,64 (17,98) | 37,09 (20,59) | 0,254 |
| MoCA | 23,64 (3,85) | 23,48 (4,20) | 23,81 (3,46) | 0,667 |
| Sit to Stand Test | 10,37 (3,49) | 9,87 (3,77) | 10,92 (3,10) | 0,131 |
| IPAQ-SF |  |  |  |  |
| HADS | 17,61 (8,31) | 17,86 (7,98) | 17,33 (8,74) | 0,752 |
| ISI | 11,34 (6,58) | 11,13 (7,21) | 11,56 (5,89) | 0,745 |
| GSES-12 | 44,66 (7,51) | 44,57 (6,49) | 44,75 (8,55) | 0,910 |
| PAM | 39,82 (6,16) | 38,92 (7,24) | 40,79 (4,61) | 0,210 |
| HLS-EUQ16 | 32,10 (7,03) | 32,94 (7,84) | 31,18 (5,98) | 0,125 |

Significant differences ( $p \leq 0.05$ ) are highlighted in bold. \*A total of thirty symptoms were included. Chi-Square test for the variables gender, marital status, educational level and occupation, and one-way ANOVA for the rest of the variables. TWD, temporary work disability; SF-36, Short Form-36 Health Survey Questionnaire; SF-36 Physical Health= physical function + physical role + bodily pain + general health; SF-36 Mental Health= vitality + social function + emotional role + mental health; MoCA, Montreal Cognitive Assessment; HADS, Hospital Anxiety and Depression Scale; ISI, Insomnia Severity Index; IPAQ-SF, Physical Activity Questionnaire-Short Form; GSES-12, Self-efficacy Scale-12; PAM, Patient Activation Measure Questionnaire; HLS-EUQ16, Health Literacy Europe Questionnaire.

When analyzing the use of ReCOVery APP in the intervention group, the range of use during the 6 months ranged from 10.95 min to 5,764.81 min. The mean use was 839.65 min (SD: 1,090.57) during the 6 months. Only 7 participants (13.4%) in the intervention group made significant use of the APP. More than 90% used the APP during the first three months, but not afterwards, which explains the low adherence in the medium term.

Table 2 shows a comparison between the intervention group and the control group, using data obtained from three different measurements, from the beginning to the end of the intervention. As shown, no significant differences in favour of the intervention group were identified between the end (T2) and the beginning of the intervention (T0). However, the SF-36 Physical Health variable did show statistically significant differences between the intervention and control groups at three months (p: 0.021; CI: −16.30, −1.40) and six months (p: 0.022; CI: −2.18, −3.18), with better levels of physical health in favour of the control group. The intervention group has improved (T2-T0) in the variables: mental health, number of persistent symptoms, cognitive status, emotional well-being, habitual physical activity, activation and health literacy, compared to the control group.

**Table 2:** Outcome data at baseline, three and six month follow-up, and their comparting by assigned groups.

| VARIABLES | INTERVENTION GROUP*<br>N (%) Mean (SD) | CONTROL GROUP**<br>N (%) Mean (SD) | SIGNIFICANCE<br>p-value (CI) |
| --- | --- | --- | --- |
| **PRIMARY OUTCOMES** |  |  |  |
| SF-36 Physical Health |  |  |  |
| Baseline (T0) | 29.06 (13.67) | 35.58 (18.86) | 0.053 (-13.12; 0.07) |
| 3 months (T1) | 33.80 (12.19) | 42.30 (20.31) | **0.021 (-16.30; -1.40)** |
| 6 months (T2) | 32.53 (18.11) | 41.88 (22.40) | **0.022 (-2.18;-3.18)** |
| T2-T0 | 5.33 (14.03) | 7.50 (17.37) | **0.106 (-9.16;4.84)** |
| SF-36 Mental health |  |  |  |
| Baseline (T0) | 32.64 (17.98) | 37.09 (20.59) | 0.252 (-12.16; 3.24) |
| 3 months (T1) | 37.35 (20.01) | 40.29 (19.59) | 0.491 (-11.38; 5.50) |
| 6 months (T2) | 44.45 (24.07) | 48.57 (24.30) | 0.845 (-14.89;6.66) |
| T2-T0 | 14.48 (19.85) | 11.87 (19.92) | 0.902 (-6.25;11.48) |
| **SECONDARY OUTCOMES** |  |  |  |
| Number of persistent symptoms |  |  |  |
| Baseline (T0) | 17.50 (5.29) | 15.35 (6.55) | 0.074 (-0.23; 4.52) |
| 3 months (T1) | 16.48 (4.65) | 14.00 (6.64) | 0.090 (-0.44; 5.41) |
| 6 months (T2) | 13.92 (6.40) | 12.13 (7.06) | 0.318 (-1.19; 4.79) |
| T2-T0 | -0.84 (3.18) | -1.67 (3.80) | 0.485 (-0.98; 2.66) |
| MoCA |  |  |  |
| Baseline (T0) | 23.48 (4.20) | 23.81 (3.46) | 0.667 (-1.85; 1.19) |
| 3 months (T1) | 24.13 (4.45) | 24.14 (3.84) | 0.991 (-1.78; 1.76) |
| 6 months (T2) | 25.48 (2.99) | 25.39 (2.98) | 0.773 (-1.37; 1.53) |
| T2-T0 | 1.00 (4.67) | 0.11 (2.87) | 0.249 (-0.86; 2.65) |
| Sit to Stand Test |  |  |  |
| Baseline (T0) | 9.87 (3.77) | 10.92 (3.10) | 0.131 (-2.42; 0.31) |
| 3 months (T1) | 10.65 (3.66) | 11.28 (3.89) | 0.462 (-2.30; 1.05) |
| 6 months (T2) | 11.00 (4.05) | 12.60 (4.77) | 0.269 (-3.62; 0.42) |
| T2-T0 | 0.48 (3.23) | 0.21 (2.28) | 0.390 (-1.04; 1.57) |
| IPAQ-SF |  |  |  |
| Baseline (T0) | 341.04 (443.87) | 308.68 (233.48) | 0.325 (-127.59;192.32) |
| 3 months (T1) | 344.09 (338.65) | 303.76 (244.52) | 0.325 (-129.72;212.42) |
| 6 months (T2) | 355.48 (491.631) | 271.32 (480.38) | 0.853 (-132.61-300.93) |
| T2-T0 | -224.89 (485.61) | -194.13 (51.47) | 0.813 (-218.51;157.12) |
| HADS |  |  |  |
| Baseline (T0) | 17.86 (7.98) | 17.33 (8.74) | 0.752 (-2.80; 3.86) |
| 3 months (T1) | 17.20 (8.72) | 16.00 (9.95) | 0.553 (-2.80; 5.20) |
| 6 months (T2) | 15.42 (9.18) | 16.90 (8.62) | 0.398 (-2.48; 5.44) |
| T2-T0 | -1.09 (4.96) | -0.97 (6.28) | 0.489 (-2.65; 2.40) |
| ISI |  |  |  |
| Baseline (T0) | 11.13 (7.21) | 11.56 (5.89) | 0.745 (-3.03; 2.17) |
| 3 months (T1) | 10.50 (5.53) | 10.33 (5.94) | 0.893 (-2.30; 2.63) |
| 6 months (T2) | 13.69 (7.47) | 11.18 (7.23) | 0.901 (-0.77; 5.78) |
| T2-T0 | -0.95 (5.52) | -1.65 (6.10) | 0.340 (-1.91; 3.33) |
| GSES-12 |  |  |  |
| Baseline (T0) | 44.57 (6.49) | 44.75 (8.55) | 0.910 (-3.21; 2.86) |
| 3 months (T1) | 43.31 (9.10) | 44.92 (8.69) | 0.399 (-5.41; 2.17) |
| 6 months (T2) | 43.14 (8.48) | 43.74 (8.21) | 0.915 (-4.31; 3.12) |
| T2-T0 | -0.85 (6.67) | 0.44 (6.06) | 0.265 (-4.16; 1.56) |
| PAM |  |  |  |
| Baseline (T0) | 38.92 (7.24) | 40.79 (4.61) | 0.210 (-4.26; 0.52) |
| 3 months (T1) | 40.24 (6.90) | 39.92 (5.72) | 0.816 (-2.39; 3.02) |
| 6 months (T2) | 40.17 (6.17) | 40.79 (4.76) | 0.101 (-3.09; 1.85) |
| T2-T0 | 18.26 (53.25) | 17.97 (50.81) | 0.906 (-23.23;23.83) |
| HLS-EUQ16 |  |  |  |
| Baseline (T0) | 32.94 (7.84) | 31.18 (5.98) | 0.125 (-1.00; 4.51) |
| 3 months (T1) | 32.00 (7.35) | 30.32 (7.16) | 0.291 (-1.46; 4.81) |
| 6 months (T2) | 30.00 (6.34) | 32.79 (8.24) | 0.767 (-0.51;6.08) |
| T2-T0 | -0.84 (3.18) | -1.67 (3.80) | 0.485 (-0.98;2.66) |

Significant differences ( $p \leq 0.05$ ) are highlighted in bold. \*Intervention group had 52 participants in T0, 47 participants in T1 and 42 participants in T2. \*\*Control group had 48 participants in T0, 44 participants in T1 and 38 participants in T2. SF-36, Short Form-36 Health Survey Questionnaire; MoCA, Montreal Cognitive Assessment; HADS, Hospital Anxiety and Depression Scale; ISI, Insomnia Severity Index; IPAQ-SF, Physical Activity Questionnaire-Short Form; GSES-12, Self-efficacy Scale-12; PAM, Patient Activation Measure Questionnaire; HLS-EUQ16, Health Literacy Europe Questionnaire.

Table 3 shows the evolution of both the control and intervention groups in the different quality and health constructs analysed after a six-month follow-up period. All participants significantly improved in physical health, mental health, number of persistent symptoms, cognitive status, physical performance, as well as levels of depression and anxiety (p<0.001).

**Table 3.** General description of the evolution of all study participants who were evaluated in the three measurements in a period of six months.

| VARIABLES | RESULTS T0<br>N=80<br>Mean (SD) | RESULTS T2<br>N=80<br>Mean (SD) | SIGNIFICANCE<br>p-value | Confidence interval 95% |  |
| --- | --- | --- | --- | --- | --- |
|  |  |  |  | Inf. | Sup. |
| **PRIMARY OUTCOMES** |  |  |  |  |  |
| SF-36 Physical health | 30,61 (14.401) | 36,97 (20.679) | **p < 0.001** | -9.847 | -2.883 |
| SF-36 Mental health | 33,16 (18.352) | 46,40 (24.120) | **p < 0.001** | -17.652 | -8.836 |
| **SECONDARY OUTCOMES** |  |  |  |  |  |
| N. ° of persistent symptoms | 16,81 (6.084) | 12,83 (6.649) | **p < 0.001** | 2.80 | 5.148 |
| MoCA | 23,59 (3.946) | 25,48 (3.241) | **p < 0.001** | -2.563 | -1.215 |
| Si-to-Stand Test | 10,22 (3.606) | 11,74 (4.446) | **p < 0.001** | -2.311 | -0.715 |
| IPAQ-SF | 326,43 (361.277) | 315,50 (485.089) | 0.805 | -76.960 | 98.835 |
| HADS | 18,26 (8.316) | 16,20 (8.868) | **p < 0.001** | 0.980 | 3.144 |
| ISI | 12,02 (6.408) | 12,05 (7.424) | 0.475 | -1.792 | 0.842 |
| GSES-12 | 43,70 (7.925) | 43,43 (8.306) | 0.703 | -1.154 | 1.705 |
| PAM | 39,75 (6.327) | 31,46 (5.523) | 0.320 | -2.129 | 0-704 |
| HLS-EUQ16 | 31,98 (7.299) | 31,46 (7.490) | 0.426 | -0.781 | 1.831 |

Significant differences ( $p \leq 0.05$ ) are highlighted in bold. SF-36, Short Form-36 Health Survey Questionnaire; MoCA, Montreal Cognitive Assessment; HADS, Hospital Anxiety and Depression Scale; ISI, Insomnia Severity Index; IPAQ-SF, Physical Activity Questionnaire-Short Form; GSES-12, Self-efficacy Scale-12; PAM, Patient Activation Measure Questionnaire; HLS-EUQ16, Health Literacy Europe Questionnaire.

Table 4 shows the regression models in relation to improvements in quality of life and mental health in both the control and intervention groups. No significant results were observed to help explain the improvement in physical health (SF-36 physical health). Results showed how increases in quality of life and mental health were predicted by an increase in time spent using the ReCOVery APP (p:0.009; p:0.003, respectively) and personal self-efficacy constructs (GSES-12) (p:0.025, p:0.012, respectively), explaining 17.8% of the variance in quality of life and 20.4% in mental health.

**Table 4:** Linear regression model in relation to the improvements of quality of life and mental health.

| Quality of life (SF-36 physical and mental health) | Coefficient | P-Value | Confidence interval 95% |  | Collinearity analysis |  |
| --- | --- | --- | --- | --- | --- | --- |
|  |  |  | SUPERIOR | INFERIOR | TOLERANCE | VIF |
| Minutes of ReCOVery APP use | 0.001 | **0.009** | 0.000 | 0.000 | 0.920 | 1.088 |
| Increase of in self-efficacy (GSES-12) | 0.675 | **0.025** | 0.088 | 1.262 | 0.879 | 1.138 |
| R2 | 0.178 |  |  |  |  |  |
| R2adj | 0.134 |  |  |  |  |  |
| Mental Health (SF-35 Mental health) | Coefficient | P-value | CI 95% |  | Collinearity analysis |  |
|  |  |  | SUPERIOR | INFERIOR | TOLERANCE | VIF |
| Minutes of ReCOVery APP use | 0.001 | **0.003** | 0.000 | 0.000 | 0.920 | 1.088 |
| Increase of in self-efficacy (GSES-12) | 0.860 | **0.012** | 0.194 | 1.527 | 0.879 | 1.138 |
| R2 | 0.204 |  |  |  |  |  |
| R2adj | 0.162 |  |  |  |  |  |

Significant differences ( $p \leq 0.05$ ) are highlighted in bold. SF-36, Short Form-36 Health Survey Questionnaire; GSES-12, Self-efficacy Scale-12.

## DISCUSION

For the scientific community, understanding the aetiology of Long COVID is proving much more complex than in the case of COVID-19 (43). Research has created evidence, but there are still many unknowns surrounding the disease. The disparity of symptoms, coupled with the presence of comorbidities, has created great difficulty in understanding the condition (44). In the last two years, several medical guidelines have been published focusing on the diagnosis and management of Long COVID (22–24). However, these guidelines lack potentially useful rehabilitation interventions, which is understandable given the paucity of RCTs conducted in this patient group. In the same vein, official sources highlight the need for interventions based on a personalised care plan, from a multidisciplinary perspective (45). In this context and given that Long COVID patients have experienced a reduction in their quality of life, functional capacities, and well-being (46), this intervention has designed ReCOVery, an APP with a multidisciplinary rehabilitative approach, focused on improving the quality of life of patients with persistent COVID.

The results of this study concluded that ReCOVery APP was not significantly more effective in improving the quality of life of patients with Long COVID over a six-month period. Effectiveness analyses have revealed a greater improvement in the intervention group compared to the control group in terms of their mental health (SF-35 mental health and HADS), cognitive ability (MoCA), physical ability (IPAQ-SF), activation ability (PAM) and health literacy (HLS-EUQ16), as well as a greater decrease in the number of persistent symptoms. However, these were non-significant improvements compared to the control group. However, linear regression analyses have revealed significant patterns of improvement in the quality of life of these patients predicted by time of APP use and self-efficacy (GSES-12).

It was from 2021 onwards that the first telerehabilitation and digital health interventions began to appear, aimed at the recovery of post-COVID-19 and persistent COVID patients. An example of this was the pilot study by Fowler-Davis et al. (2021), which identified improvements in quality of life, including through a virtual consultation to offer advice on their needs, albeit with only eight participants (47). In contrast, the telerehabilitation programme of Dalbosco-Salas et al. (2021) had a large number of Post-COVID-19 participants with persistent dyspnoea, although there was no control group. This intervention was based on 24 supervised home-based exercise training sessions for patients and achieved significant improvements in fatigue, dyspnoea, physical function and quality of life as measured by the SF-36. However, they did not report improvements in their physical complaints, nor in their perceived mental health through telerehabilitation (48). Harenwall et al (2021) conducted an intervention based on a course of recovery after COVID infection, and reported an increase in their general health after the intervention. It appears that the inclusion criteria allowed for self-diagnosis by self-suspicion of the participants and no inclusion criteria were set so that the sample has low validity (49). Thus, early research would point towards telerehabilitation as a viable option for the recovery of patients with Long COVID, but had several of the biases already mentioned.

In 2022, the first RCTs started to be implemented, such as the one by Li et al. (2022), based on a 6-week unsupervised home-based post-COVID-19 exercise programme. This study did obtain significant improvements in favour of the intervention group in their quality of life (SF-12), although both groups improved lung function in a similar way but no long-term effects were obtained (50). The RCT by McNarry et al (2022) verified that after eight weeks of inspiratory muscle training in Post-COVID-19 patients there was no improvement in quality of life in favour of the intervention group (51). The cohort study by Kortianou et al. (2022) studied the clinical effects of a home-based telerehabilitation exercise programme after discharge from hospital for COVID-19. Their results showed significant improvements in perceptions of health and well-being (measured through the SF-36 and HADS), however, they did not show significant improvements in physical health. In addition, more than half of their participants dropped out of the intervention before completion (52). On the other hand, the clinical trial by Estébanez-Pérez achieved a significant improvement in the functional capacity of patients with Long COVID after a 4-week digital physiotherapy intervention. However, this study did not have a control group of participants (53). Finally, the qualitative study by Ruckser-Scherb et al. (2022) analysed the experiences of post-COVID-19 patients after two weeks of using an APP, initially for oncology patients (54). In response, more than 80 % of participants found some of the content useful, more than 70 % liked the exercises, and all participants recommended its use to post-COVID-19 and persistent COVID patients.

Regarding the most recent evidence, the systematic review by Chuang et al. (2023) affirm that therapies based on physical exercise are fundamental for the recovery of Long COVID, including the telerehabilitation modality (55). In addition, I recommend early management that should begin with a comprehensive and individualized evaluation (55). In this sense, De Mars et al. (2023) publish recommendations to implement a safe rehabilitation for this group of patients (56). Among them, the RCT by Del Corral et al. (2023), in which, through a supervised 8-week inspiratory/expiratory muscle program carried out from home, he managed to improve the quality of life of the 88 post-COVID-19 participants. No improvements in exercise tolerance were achieved (57). The RCT by Santana et al. (2023) investigated the potential therapeutic effects of high-definition transcranial direct current stimulation in conjunction with a rehabilitation program for the management of post-COVID-19 fatigue. This intervention was effective in reducing fatigue and anxiety, as well as improving quality of life (58). The Jimeno-Almazán RCT demonstrated the benefits of a supervised exercise program in people with post-COVID-19 conditions, for people with mild COVID-19 (59). In this line, the recent systematic review by Burnett et al. (2023) show promise for exercise interventions in Long COVID patients for improving functional exercise capacity, dyspnea, and fatigue (60). The scoping review by Rinn et al. (2023) believe that digital interventions have the potential to help control some persistent symptoms, both physical and psychological (61).

Thus, the use of digital interventions, such as telerehabilitation, is being progressively implemented for a multitude of pathologies, and reported research suggests that this is also the case for Long COVID (61). Current evidence demonstrates the potential of digital interventions to help control some physical and psychological symptoms. Several studies on telerehabilitation that evaluated quality of life obtained significant improvements in favor of the intervention group (62–65). However, its inclusion criteria allowed access to patients infected for less than twelve weeks, so it could not be considered that the total sample was made up of Long COVID patients, as some indicate. Thus, the limited research on effective digital tools for patients with Long COVID still requires the implementation of large-scale RCTs with a high degree of clinical evidence. An important aspect to consider in future research has turned out to be adherence. Various investigations have reported numerous dropouts of a notable percentage of participants (48,52,62), as well as poor adherence as shown in this study. However, in the short term, the time of use of ReCOVery APP was not found to be correlated with the improvement of the general quality of life and mental health of the participants (26). Therefore, these medium-term analyzes are promising given the effectiveness of APP.

Of particular note is the construct of self-efficacy, which is also correlated with better overall quality of life and mental health. Self-efficacy is a factor with potential for inclusion in the ability of patients with chronic diseases to self-manage their symptoms (66). Thus, patients with high self-efficacy in coping with their chronic diseases show more perceived ability to manage the challenges related to their diseases and a greater sense of control over their lives (67). This evidence would be consistent with the findings that higher self-efficacy correlates with better overall quality of life and mental health. This reality has also been seen in other pathologies such as multiple sclerosis (68), oncological processes (67) or after the sequelae of a stroke (69). Therefore, the role of self-efficacy should be considered as an important element in the recovery of patients with persistent COVID.

Finally, participants in the present intervention did not report any unanticipated adverse effects, either during the intervention or months later. Furthermore, there were no difficulties of any kind during the course of the intervention. Therefore, it can be affirmed that this is a safe, consistent and feasible intervention. Our research team intends to carry out a qualitative analysis to further investigate the low adherence of the participants, as well as to be able to offer technical and content improvements in the future.

Our study has some limitations. First, the context in which the study was carried out, during the vaccination process and an increase in the number of infections. Second, due to the nature of the intervention, all participants were aware of their group assignment during the RCT. Third, the symptomatology itself (physical and mental) and its fluctuation may be limiting for rehabilitation interventions. Fourth, the Montreal Cognitive Assessment (MoCA) questionnaire obtained a Cronbach’s alpha of 0.49, thus reporting low reliability. Finally, the study variables are based on participants’ self-perceptions; therefore, we must rely on their statements, even if they cannot be objectively verified. This study also has several strengths. Unlike other research, a specific APP has been designed for persistent COVID, which is easy to use and access from any location with an internet connection. In addition, to our knowledge, there are no studies in our country with a similar number of Long COVID participants and a long follow-up time, using a telerehabilitation intervention for this disease.

## CONCLUSIONS

After a six-month telerehabilitation intervention for patients with persistent COVID, some non-significant improvements were observed. However, linear regression analyses revealed that longer use of ReCOVery APP and higher self-efficacy lead to better overall quality of life and mental health of these patients. Possibly, low adherence to the APP justifies the lack of significant improvements. Therefore, future large-scale research is needed to further investigate strategies to increase adherence to telerehabilitation in patients with persistent COVID. Ultimately, the development of this research increases the knowledge available on the effectiveness of telerehabilitation for patients with persistent COVID as a promising tool to improve their quality of life.

## Data Availability

The data sets used
and/or analyzed during this study are available on the ZENODO platform. They were registered and published on 02/20/2023 with DOI: 10.5281/
zenodo.7656949.

## DECLARATIONS

### Acknowledgements

We wish to thank the University of Zaragoza, the Aragonese Primary Care Research Group (GAIAP, B21_23R) that is part of the Department of Innovation, Research and University at the Government of Aragón (Spain); the Institute for Health Research Aragón (IIS Aragón); the Research Network on Chronicity, Primary Care, and Health Promotion (RICAPPS) that received a research grant from the Carlos III Institute of Health, Ministry of Science and Innovation (Spain), awarded on the call for the creation of Health Outcomes-Oriented Cooperative Research Networks (RICORS), with reference RD21/0016/0005, co-funded with European Union – NextGenerationEU funds, which finance the actions of The Recovery and Resilience Facility (RRF); the University of Zaragoza; and Feder Funds “Another way to make Europe”.

### Authors’ contributions

M.S.-P., B.O.-B. and S.L.-H. elaborated the research design. M.S.-P., B.O.-B. and S.L.-H. Developed the study and coordinated the field work. R.S.-R and M.S.-P did the analysis. R.S.-A., V.C.-V and R.S.-R. have helped with the coordination of the project. M.S.-P. and B.O.-B. wrote the manuscript. B.O.-B. is the principal investigator of the project. All authors reviewed the content of the manuscript and approved the final version for submission. Not applicable as the data are anonymised and no individual images are presented.

